# LUTS-V: A New Simplified Score for Assessing Lower Urinary Tract Symptoms in Men

**DOI:** 10.1101/2020.05.05.20091835

**Authors:** Caroline Santos Silva, Ueslei Menezes de Araujo, Mateus Andrade Alvaia, Kátia Santana Freitas, Cristiano Mendes Gomes, José de Bessa Júnior

## Abstract

**OBJECTIVE:** To validate a new simplified score for the assessment of men with LUTS (LUTS-V).

**METHODS:** We made adjustments to the VPSS, resulting in a new simplified instrument (LUTS visual score – LUTS-V). In a pilot study, LUTS-V was administered to 50 men to identify interpretation issues. We used the International Prostate Symptom Score (IPSS) as the gold standard to validate the new tool in 306 men. The total IPSS and LUTS-V scores for each subject were evaluated and we used Bland-Altman analysis and Pearson’s correlation plot to assess the agreement between the scores. A ROC curve was utilized to determine the diagnostic accuracy of LUTS-V and its diagnostic properties were described in terms of sensitivity, specificity, positive and negative predictive values.

**RESULTS:** Median age was 59 [52-67] years and, according to the IPSS, 26 (8.7%) patients had severe symptoms, while 99 (33%) had moderate symptoms, and 175 (58.3%) had mild symptoms. We found a positive correlation between the IPSS and LUTS-V (r = 0.72; p < 0.0001). Bland-Altman analysis showed good agreement between the two questionnaires. We found LUTS-V to have a diagnostic accuracy to detect more severe cases of 83% (95% CI: [78-87%]; p < 0.001), as estimated by the area under the ROC curve. The cut-off value of ≥ 4 points was the best threshold, with a sensitivity of 74% and a specificity of 78%, which resulted in a negative predictive value of 81% and a positive predictive value of 71% in this scenario. Median completion time was 0.51 [0.41-1.07] min for LUTS-V and 2.5 [2.2-3.4] min for the IPSS (p < 0.0001). In addition, 91.5% of patients completed the questionnaires with no help, while the other 8.5% were interviewed.

**CONCLUSION:** LUTS-V is a simple, self-administered tool with a significant discriminating power to identify patients with moderate to severe symptoms.

## INTRODUCTION

Lower urinary tract symptoms (LUTS) comprise a variety of urinary symptoms and are very common among adult men(1,2).They can be detrimental to the quality of life of affected individuals and are frequently associated with other clinical conditions such as diabetes, neurological disorders, and erectile dysfunction(3,4).Because they are quite common, have a negative impact upon quality of life and often warrant a diagnostic workup and treatment, LUTS can lead to increased costs to both the individual and the community(5,6).

The assessment of men with LUTS must be focused and take into account all aspects that might be relevant to the differential diagnosis, enabling the clinician to identify symptom severity and associated bother as well as recognizing those who require a more thorough evaluation(7).

Different guidelines recommend the use of a validated symptom questionnaire in the initial evaluation of men with LUTS(7,8). Patient-reported outcome assessments are considered effective tools for characterizing symptom burden and health-related quality of life, and they are playing an increasing role in clinical decision-making(9). The International Prostate Symptom Score (IPSS) is the most widely used questionnaire for the evaluation of men with LUTS(10). An additional question evaluates the impact of LUTS on quality of life(11) It stratifies patients in terms of symptom severity and may be used to monitor disease progression and response to treatment(12).

The use of patient-reported outcome measures may be limited by their extension or the complexity of their questions and response options. Ideally, they should be as short as possible, enabling easy and rapid completion, which may help expand their usage and improve their accuracy(13).

Patients with a low education level have been demonstrated to have difficulty completing the IPSS accurately. The difficulty in understanding the IPSS questions, even for men with a relatively high education level, often leads patients to ask for help when completing the questionnaire. This introduces the risk of unwarranted interference in patient responses (14,15). The use of simplified, more accessible instruments has been proposed to ease questionnaire completion and minimize interference (16,17).

The Visual Prostate Symptom Score (VPSS) created by van der Walt and colleagues comprises pictograms designed to evaluate three of the seven symptoms evaluated in the IPSS: urinary frequency, nocturia, weak stream and also their impact on quality of life. The VPSS significantly correlates with IPSS and can be completed with no assistance by a greater proportion of men with limited education, indicating it may be more useful than the IPSS for illiterate patients or men with a low education level (16).

Despite its improved applicability (18), the VPSS present some limitations. According to a 2016 study, items that evaluate nocturia and quality of life were deemed unclear by many participants and the pictogram dark background was also significantly criticized. Suggested improvements included use of larger image for the pictograms depicting urinary frequency and nocturia and the inclusion of images depicting urinary urgency (19). Although not previously highlighted, we found additional limitations including the lack of an option for nocturia zero times, the fact that normal values for daytime urinary frequency such as 4 micturitions/day are scored as increased frequency, the difficulty in interpreting the micturating flow and the use of multiple pictograms for quality of life, resulting in a “ceiling and floor” effect.

The aim of this study was to validate a new simplified visual score for the assessment of men with LUTS, which was inspired from the experience with the VPSS.

## METHODS

By modifying the Visual Prostate Symptom Score (VPSS) (16), we developed a new visual score (LUTS-V - Figure 1). From the VPSS settings, we made the following changes in this new version: a) changes in the images and the sequence; b) inclusion of new response options based on the associated deficiencies; c) reduction in the number of options for answering a question about the quality of life from 7 to 3 options. The authors reviewed the LUTS-V score to ensure the original content of the VPSS had been maintained and to detect inconsistencies with the new version. The results were submitted to a committee composed of two urologists specialized in voiding dysfunction, one physical therapist and one nurse.

**Figure 1:**
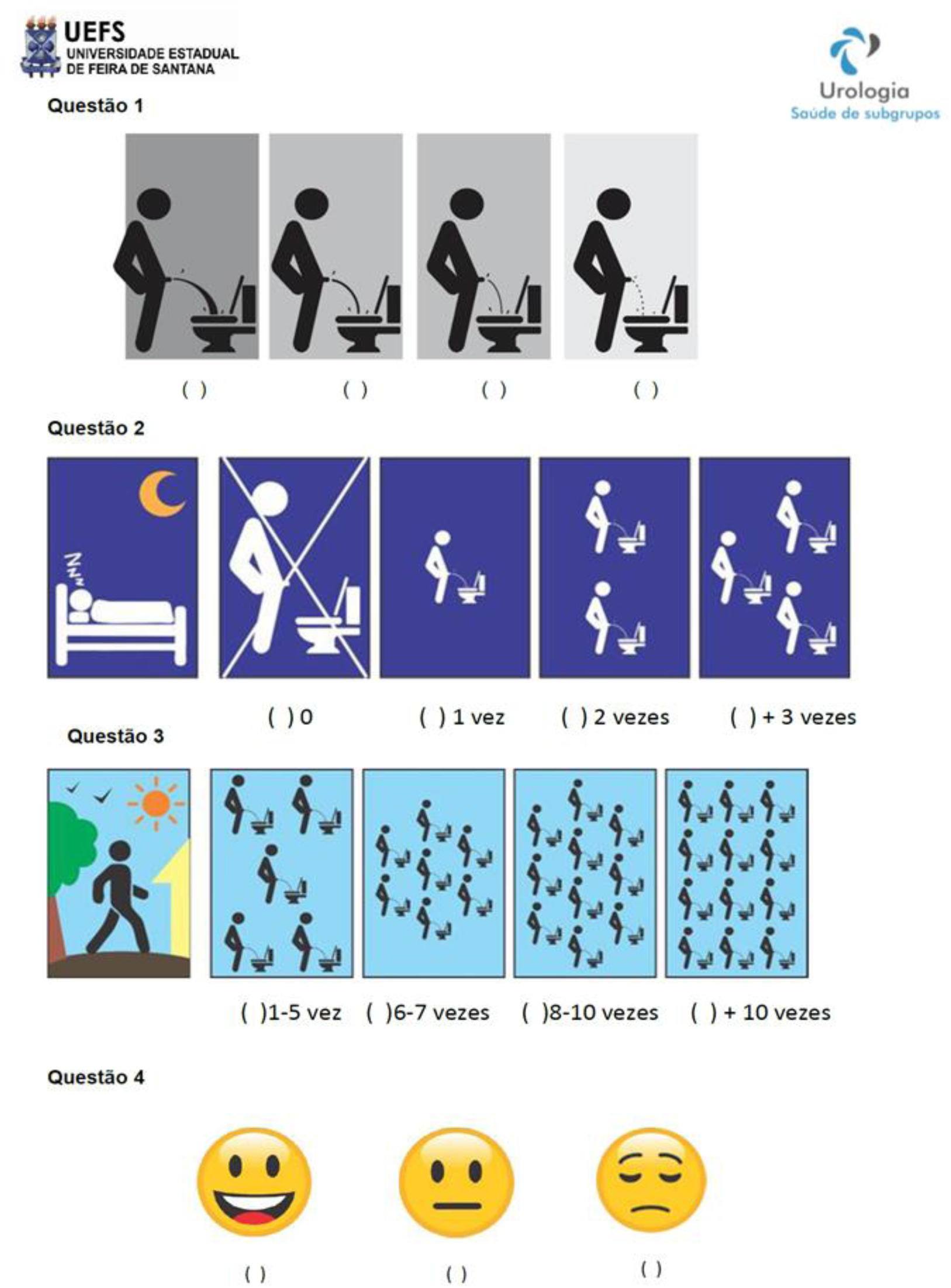
LUTS-V final version

In a pilot study, LUTS-V was administered to 50 men aged > 40 years. Respondents were asked about their understanding of the questions and whether the response options were clear. After minor final adjustments made by the committee based on the responses obtained in this pilot study, the test version of the scale was concluded.

Men older than 40 years who presented to a urological outpatient unit met our inclusion criteria. We used the following exclusion criteria: a history of urological surgery in the past 12 months, an acute change in general health status during follow-up, and patients with cognitive impairment.

The study cohort consisted of consecutive men who attended urologist office visits between January 2018 and June 2018. Participants were asked to complete both the LUTS-V and the IPSS surveys at baseline.

The questionnaires were self-administered in a private and quiet room. Patients were allowed to ask for the assistance of a designated researcher in case of difficulty understanding or completing the questionnaires. Illiterate men completed the questionnaires in the form of a structured interview. After completing LUTS-V, all participants were asked if they had understood each of the items and had found suitable answers. In the event of lack of comprehension of any item or difficulty in identifying a suitable answer, the reason given by the patient was noted by the researcher in an appropriate file.

The COSMIN (Consensus-based Standards for the Selection of Health Status Measurement Instruments) guidelines were used to guide analysis and reporting(20). This study was approved by the Research Ethics Committee of the State University of Feira de Santana under the protocol no. 64704017.7.0000.0053, position statement 2.052.761 (ANNEX D), and all participants provided written informed consent.

Data were expressed as medians and interquartile ranges, or absolute values and fractions. The Mann-Whitney U test were used to compare continuous variables, while the chisquared and Fisher’s exact test were used to compare categorical variables.

Both the IPSS (score of 0 to 7 indicates mild symptoms, 8 to 19 indicates moderate symptoms, and 20 to 35 indicates severe symptoms) and the LUTS-V (range: 0-11 points) surveys were used as data collection instruments; the former was considered the gold standard. The total IPSS and LUTS-V scores for each subject were used to determine the agreement between the two instruments using Bland-Altman analysis and Pearson’s correlation plot.

A ROC curve was used to evaluate the diagnostic accuracy and the best cut-off point for LUTS-V. Diagnostic properties (content validity) were described in terms of sensitivity, specificity, and diagnostic odds ratios.

Uroflowmetry was used as a reference standard for the construct validity analysis of LUTS-V through hypothesis testing and to determine the maximum urinary flow (Qmax). We expected the urinary flow rate to decrease as the total LUTS-V score increased.

ANOVA was used to compare these data and evaluate between-group differences and linear trends.

To assess the respondent burden, the time necessary for completion (in minutes) of each questionnaire (IPSS and LUTS-V) was measured and the need for assistance to complete them was noted.

All tests were two-sided, with a p < 0.05 considered statistically significant GraphPad Prism, version 8.03, were used for data analysis.

## RESULTS

The final study sample comprised 306 men aged 59 [52-67] years, 26 (8.7%) of whom had severe symptoms, while 99 (33%) had moderate symptoms, and 175 (58.3%) had mild symptoms according to the IPSS. We found a positive correlation between the IPSS and the LUTS-V total scores (r = 0.72; 95% CI: [0.65-0.77]; p < 0.0001) (Figure 2), including quality of life (r = 0.76; 95% CI: [0.69-0.83]; p < 0.0001).

**Figure 2:**
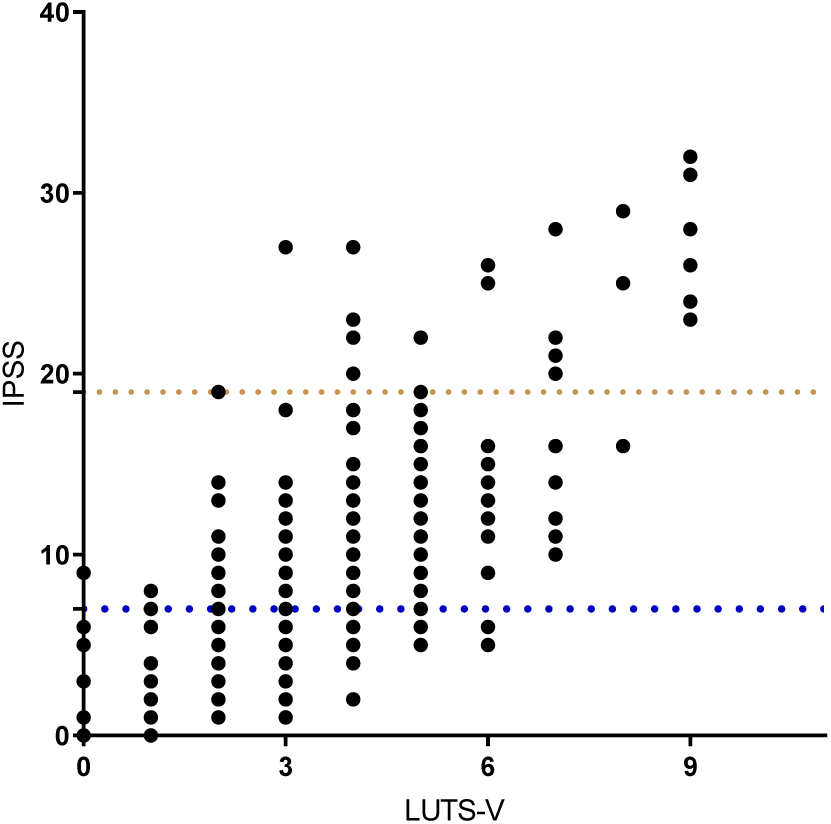
Spearman’s correlation between the IPSS and LUTS-V.

Bland-Altman analysis showed good agreement between the two questionnaires (Figure 3) (bias = 0.056; p < 0.001). Maximum urinary flow rates were found to be significantly lower in moderate and severe cases when compared to those with mild symptoms, i.e., 12 ml/s [8-18] and 17 ml/s [13-25], respectively (p < 0.001), with a median difference of 5 ml/s.

**Figure 3:**
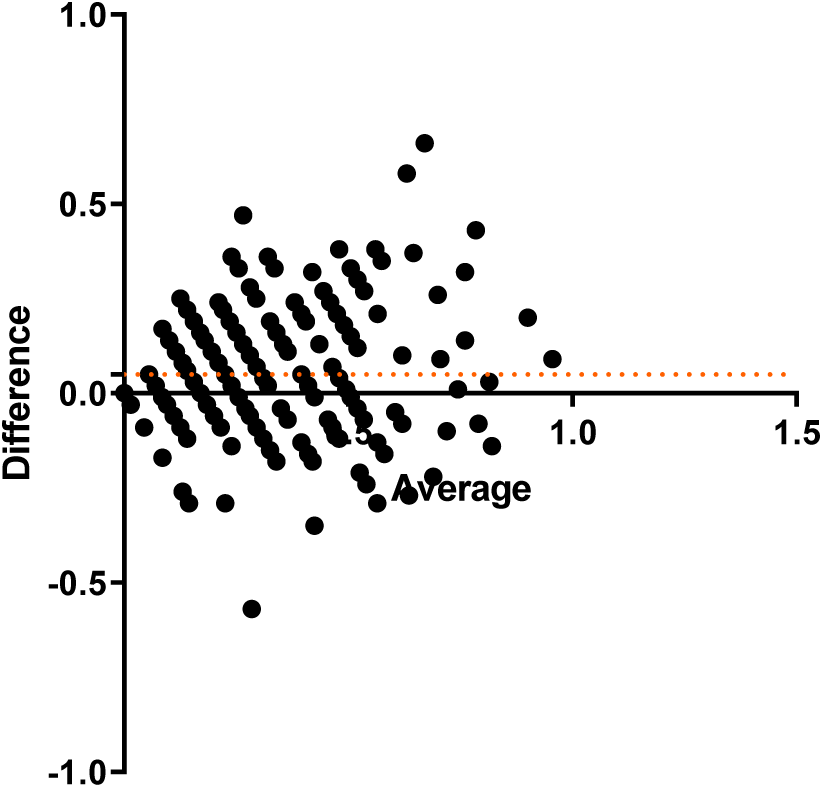
Bland-Altman plot showing agreement between the IPSS and LUTS-V.

Furthermore, maximum urinary flow rates decreased in correlation with the pictograms depicting the force of the urinary stream, with the following median Qmax values: A = 17.5 [13-16], B = 15 [11-23], C = 12 [8-18], and D = 9.3 [5.7-12.2] ml/s (A to D; p < 0.001) (Figure 4).

**Figure 4:**
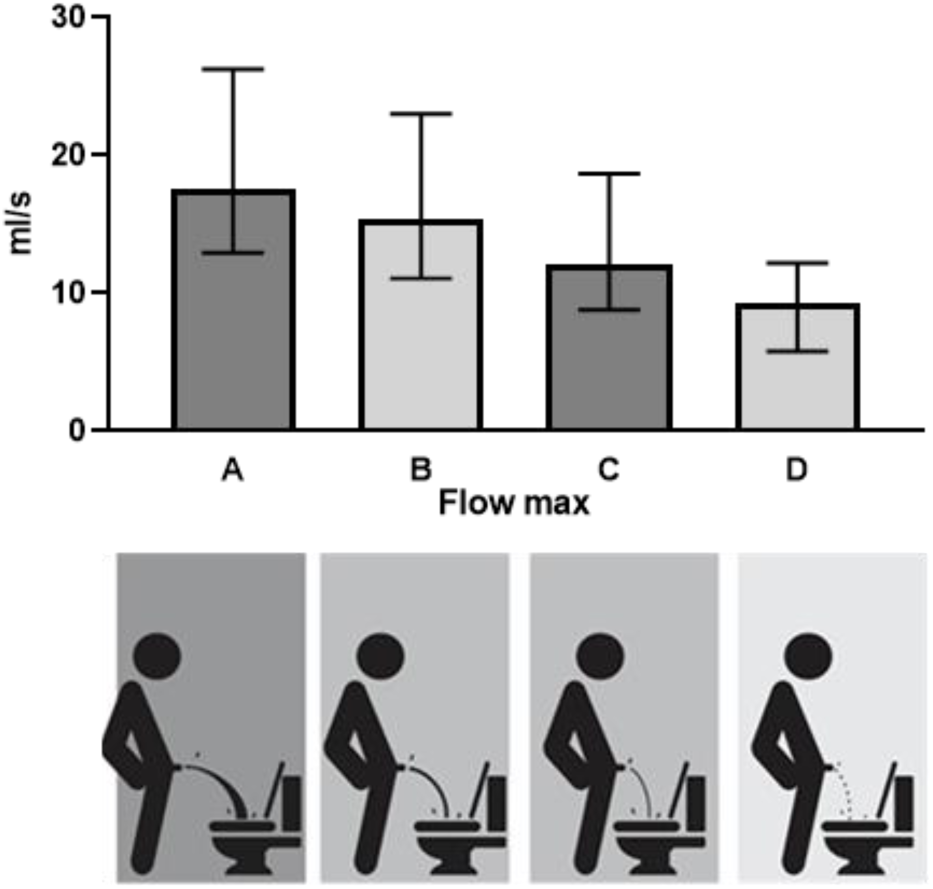
Maximum urinary flow according the urinary stream pictograms

We found LUTS-V to have excellent diagnostic accuracy to detect more severe cases, with an area under the ROC curve of 83% (95% CI: [78-87%]; p < 0.001) (Figure 5).The cut-off value of ≥ 4 points yielded a sensitivity of 74% and a specificity of 78%, which resulted in a negative predictive value of 81% and a positive predictive value of 71% in this scenario.

Median completion time was 0.51 [0.41-1.07] min for LUTS-V and 2.5 [2.2-3.4] min for the IPSS (p < 0.0001). Of the total 306 participants, 280 subjects (91.5%) completed the questionnaires without any help, while the other 26 (8.5%) were interviewed. The patients who needed assistance were significantly older (72 [62-74] versus 58 [51-64] years; p < 0.001) and had a lower education level (4 [2-7] versus 11 [8-14] years of education; p < 0.001).

**Figure 5:**
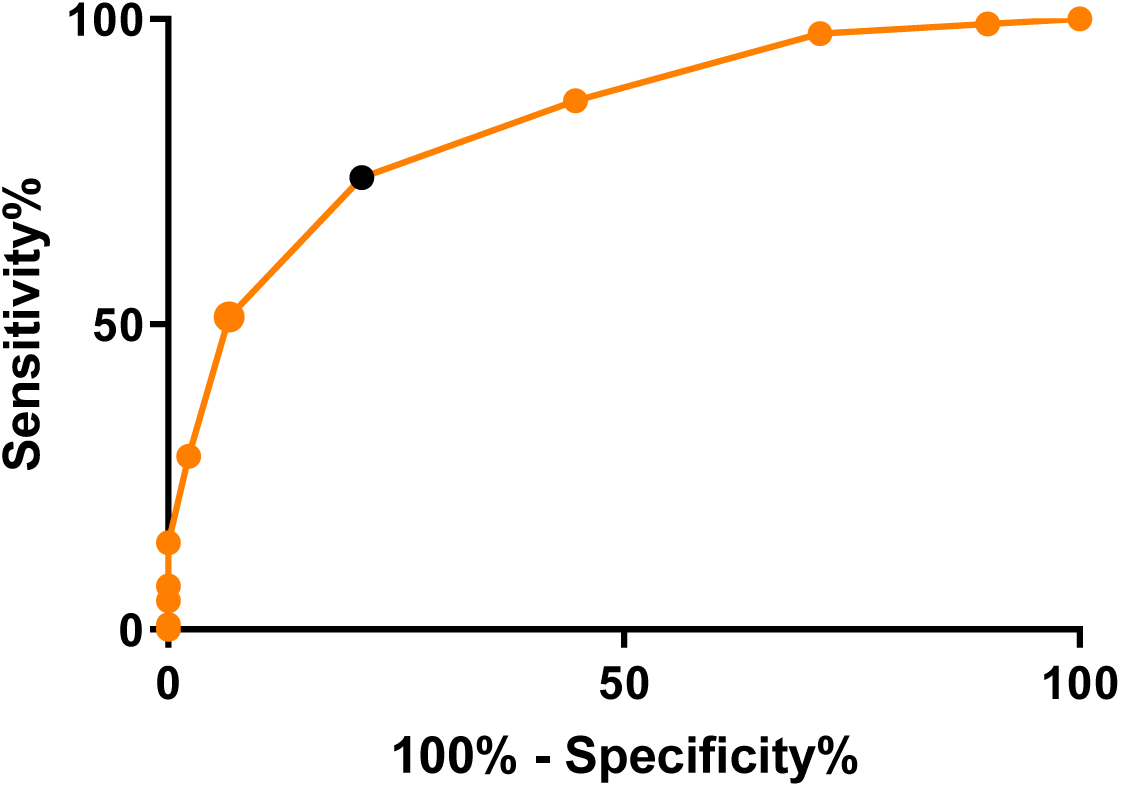
ROC curve for LUTS-V to discriminate moderate to severe cases.

## DISCUSSION

LUTS are known to be very common, especially among older patients(1,21). In the Male Attitudes Regarding Sexual Health study, carried out in 2007 in the USA, the general prevalence of LUTS was 28%, of which 40% had moderate or severe symptoms (22). According to a 2006 telephone-based survey conducted in Canada, Germany, Italy, Sweden, and the UK, the general prevalence of LUTS was 62.5%(1). A study conducted in five large Brazilian cities found the prevalence of severe LUTS among men older than 40 years to be 30% and 40% of patients were very dissatisfied with this condition (23). In the present study, we found the prevalence of moderate and severe LUTS to be 40% in the age group studied.

The VPSS is a validated instrument for the diagnosis of these symptoms, comprising four questions, and was created as a simplified survey (16) based on the International Prostate Symptom Score (IPSS) domains (10). Following the initial publications describing the development and validation of the VPSS, other studies utilized this tool to appraise its validity and applicability when compared to the gold standard, the IPSS (24,25), and urodynamic findings, namely uroflowmetry (26). Despite being widely used, the VPSS has some understanding and interpretation issues, especially common among older men and those with a lower education level (19,27).

These limitations include: normal responses for the question about daytime urinary frequency which are erroneously classified as severe; the lack of a zero value for the question about nocturia (absence of nocturia); the difficulty in understanding the image depicting the force of the urinary stream, which shows different streams at once and may be interpreted by some patients as the “urinary flow sequence”; and too many pictograms for quality of life, which may be confusing, leading patients to choose extreme response options. In this study, we therefore aimed to adapt the VPSS by correcting these problems.

Simplified questionnaires have been recommended in the clinical practice of primary care health professionals as screening instruments, particularly for patients with known risk factors, to aid in the stratification of a condition and in the subsequent investigation into the potential worsening of detrimental health issues such as erectile dysfunction (28). The use of simplified instruments to detect LUTS in primary care settings has also been reported (29,30).

Pictograms have been increasingly used among these simplified instruments. A study conducted by Descazeaud and colleagues (18), which aimed to validate a French pictogram score for evaluating LUTS, has found comparable diagnostic properties to the IPSS; however, although very similar to the VPSS, the authors included a pictogram for urinary urgency, expanding the scope of symptoms assessed in their score. In a different visual analog scale proposed by Preciado-Estrella et al.(31) and termed the GEA scale, the authors used pictograms for all the IPSS questions in order to test its applicability (completion time and need for assistance).

Our goal in creating the LUTS-V was to develop a new simplified visual score for the assessment of men with LUTS. The reasons for that were: first, to enable the use of a simplified, more accessible instrument that may be more useful than the IPSS for illiterate patients or men with a low education level. Second, to overcome the limitations of an existing visual instrument, the VPSS. In addition to being even quicker for the patient to complete, this minimizes the time a clinician would need to spend reviewing the results and scoring it.

When comparing the two instruments the LUTS-V with the IPSS, we found a strong positive correlation. A high level of agreement was demonstrated in the Bland-Altman plot, which indicates LUTS-V has diagnostic properties that are similar to those of the IPSS.

We also found strong correlations between the maximum flow rate and the pictograms depicting the force of the urinary stream which is consistent with the relationship we hypothesized would exist during development of LUTS-V. LUTS-V to have excellent diagnostic accuracy to detect more severe cases, with an area under the ROC curve of 83%.The cut-off value of ≥ 4 points yielded a sensitivity of 74% and a specificity of 78%, which resulted in a negative predictive value of 81% and a positive predictive value of 71% in this scenario. The high sensitivity and specificity yielded by LUTS-V with a cut-off score of ≥ 4 points (classified as severe) enable this instrument as a useful screening tool, as it allows for the selective referral of individuals at higher risk to specialized care according to specific guidelines(28). We also found strong correlations between the maximum flow rate and the pictograms depicting the force of the urinary stream which is consistent with the relationship we hypothesized would exist during development of LUTS-V.

The completion time for LUTS-V was much shorter than for the IPSS 91.5% of the participants were able to complete the questionnaires without any help. As expected, patients who needed assistance were older and had a lower education level.

Furthermore, we found good agreement between the quality of life as measured by the IPSS and LUTS-V. We found this to be a strong point of the LUTS-V, since we reduced the response options for QoL from 7 to 3 which is helpful to improve patient comprehension and reduce the respondent’s burden. These results are similar to those found by Crawford and colleagues(32) when developing and validating their simplified instrument UWIN (Urgency, Weak Stream, Incomplete Emptying and Nocturia), which also contains fewer response options regarding quality of life.

Uroflowmetry is the most widely used urodynamic study in clinical practice and determines the urinary flow rate over time. The maximum urinary flow rate (Qmax) is the most widely used variable to define voiding dysfunction anda Qmax < 10 ml/s in men has a positive predictive value for detecting obstruction of 88%. Our findings are in line with those of other studies validating the VPSS in relation to urodynamic data (26). In line with Rogel’s study, which validated the Analogical Uroflowmetry tool (ANUF) (33), our urinary stream pictograms were found to be directly correlated with maximum flow rates.

The LUTS-V survey was completed more quickly, who found it easier to understand. Its applicability, use of somewhat entertaining pictograms and diagnostic properties enable LUTS-V as an alternative to the IPSS and may warrant its wide implementation in primary care settings. We believe such actions would considerably benefit men’s health, particularly in the screening of more severe cases.

## CONCLUSION

LUTS-V is a simple, self-administered tool with a significant discriminating power to identify patients with moderate to severe symptoms. It may be a useful instrument for the diagnosis and follow-up of men with LUTS, particularly in primary care settings and among patients with a low education level.

## Data Availability

The database is under the care of the senior author and will be published online. currently, the website of our research group is being improved. We believe that it will be updated in 40 days

